# Misclassification of a frequent loss of function variant from *PMS2CL* pseudogene as a *PMS2* variant in Brazilian patients

**DOI:** 10.1101/2024.03.26.24304914

**Authors:** Anthony Vladimir Campos Segura, Sara Iolanda Oliveira da Silva, Karina Miranda Santiago, Rafael Canfield Brianese, Dirce Maria Carraro, Giovana Tardin Torrezan

## Abstract

*PMS2*, a Lynch Syndrome gene, presents challenges in genetic testing due to the existence of multiple pseudogenes. This study aims to describe a series of cases harboring a rare LoF variant in the *PMS2CL* pseudogene that has been incorrectly assigned to *PMS2* with different nomenclatures. We reviewed data from 647 Brazilian patients who underwent multigene genetic testing at a single center to identify those harboring the *PMS2* V1:c.2186_2187delTC or V2:c.2182_2184delACTinsG variants, allegedly located at *PMS2* exon 13. Gene-specific PCR and transcript sequencing was performed. Among the 647 individuals, 1.8% (12) carried the investigated variants, with variant allele frequencies ranging from 15 to 34%. By visually inspecting the alignments, we confirmed that both V1 and V2 represented the same variant and through gene-specific PCR and *PMS2* transcript analysis, we demonstrated that V1/V2 is actually located in the *PMS2CL* pseudogene. Genomic databases (ExAC and gnomAD) report an incidence of 2.5% - 5.3% of this variant in the African population. Currently, V1 is classified as “uncertain significance” and V2 as “conflicting” in ClinVar, with several laboratories classifying them as “pathogenic”. We identified a frequent African *PMS2CL* LoF variant in the Brazilian population that is misclassified as a *PMS2* variant. It is likely that V1/V2 have been erroneously assigned to *PMS2* in several manuscripts and by clinical laboratories, underscoring a disparity-induced matter. Considering the limitations of short-read NGS differentiating between certain regions of *PMS2* and *PMS2CL*, using complementary methodologies is imperative to provide an accurate diagnosis.

## Introduction

The use of Next-Generation Sequencing has significantly improved the accessibility and effectiveness of genetic diagnosis for patients with hereditary cancer predisposition syndromes. However, challenges arise in complex variants, such as errors found in classifying variants within genes that have homologous genomic regions or pseudogenes with sequences very similar to the original gene. These findings highlight the ongoing need for refinement in variant identification and classification methodologies.

*PMS2* is a gene involved in DNA mismatch repair. Deficiencies in this gene are associated with Lynch Syndrome (LS), a condition related to increased risks for developing colorectal, endometrial, ovarian and other cancers. Genetic testing for this gene is challenging due to the existence of multiple pseudogenes [1, 2]. Fourteen pseudogenes have been identified and described as ψ1 to ψ14, overlapping with some or all of *PMS2* exons 1 to 5 and varying in length. Additionally, there is the *PMS2CL* pseudogene (formerly known as ψ0) with high homology to the 3’ end of *PMS2* in exons 9 and 11 to 15 [3, 4].

Prior research has highlighted the misclassification of the loss-of-function (LoF) variant c.2182_2184delACTinsG within the *PMS2CL* pseudogene, mistakenly ascribed to exon 13 of the *PMS2* gene [5]. In this study, we describe a series of cases harboring this frequent LoF variant in the *PMS2CL* that has been incorrectly identified as a pathogenic variant in *PMS2*, with varying nomenclatures throughout the years. We confirmed the variant location in *PMS2CL* using different molecular techniques in several patients. Additionally, we discuss the importance of performing alternative methods to circumvent the NGS limitations in distinguishing similar regions shared between *PMS2* and its pseudogenes and the relevance of increasing sequencing efforts across diverse populations. Finally, we empathize the pivotal role these endeavors play in ensuring precision in genetic diagnoses and mitigating disparities in genetic testing.

## Methods

### Patient cohort and data collection

We collected retrospective data from 647 patients who performed genetic testing with multigene panels harboring 26 to 126 cancer predisposing genes between 2018 and 2023 at the A.C. Camargo Cancer Center. A retrospective analysis was carried out using Sophia DDM platform to identify patients with *PMS2* (NM_000535.5) c.2182_2184delACTinsG (V1) or c.2186_2187delTC (V2) variants. Clinical information (age of onset, tumor histology, familial history of cancer) was collected from hospital electronic records. All patients signed a written informed consent and were included in studies approved by the Institutional Review Board of A.C.Camargo Cancer Center (protocol numbers 2483/18 and 2497/18).

### Gene-specific PCR

Germline DNA from saliva or leucocytes from 12 patients with the presence of V1/V2 variant were obtained. A gene-specific PCR (GSP) was performed to examine exon 13 of *PMS2* and exon 4 of *PMS2CL*, like described by Hendricks [6]. Briefly, PCR primers were design to anneal preferentially in the desired gene by positioning the primer in a variable region between *PMS2* and *PMS2CL* that contains three mismatched bases between gene and pseudogene. Primers sequences used were PMS2_E13GSP_F: GAAGTTTTGTGACACTTAGCTGA<u>G</u>T<u>AG</u> and PMS2_E13GSP_R: TTGGCCTCCCAGAGTGCTG; PMS2CL_E4GSP_F: TTGTGACACTTAGCTGA<u>A</u>T<u>TA</u>TGTTGT and PMS2CL_E4GSP_R: TTATGTTAGCGAGGCTGGTCTCAAAC (underscored bases refer to those 3 discriminating bases). PCR products were analyzed by amplicon NGS using the Ion GeneStudio S5 system, followed by sequence analysis using the Integrative Genomics Viewer (IGV).

### Transcript analysis

As a complementary and confirmatory analysis, RNA was extracted from peripheral blood of 2 patients. RNA was then converted into cDNA and subjected to a nested PCR to analyze the transcription of *PMS2*. The first PCR targeted regions from exons 10 to 15, and the second PCR focused on exons 12 to 14. The PCR products were subjected to amplicon sequencing NGS, and the sequences were analyzed using the CLC Genomics workbench software.

## Results

The allegedly *PMS2* variant received two different nomenclatures on previous testing: V1: c.2186_2187delTC; p.(Leu729Glnfs) - rs587779335, and V2: c.2182_2184delinsG (p.Thr728Alafs) - rs1554294508. We observed that the nomenclature of the variant changed in 2020, after a software update. Upon visual inspection of the alignments from patients with these variants, we confirmed that both nomenclatures represent the same variant (Figure 1A). The difference between the variants is that for V1, a single nucleotide variant (SNV) *PMS2*:c.2182A>G; p.(Thr78Ala) is called as a separated variant, while in V2 this variant is considered part of the delins event, as recommended by HGVS nomenclature.

**Figure 1.**
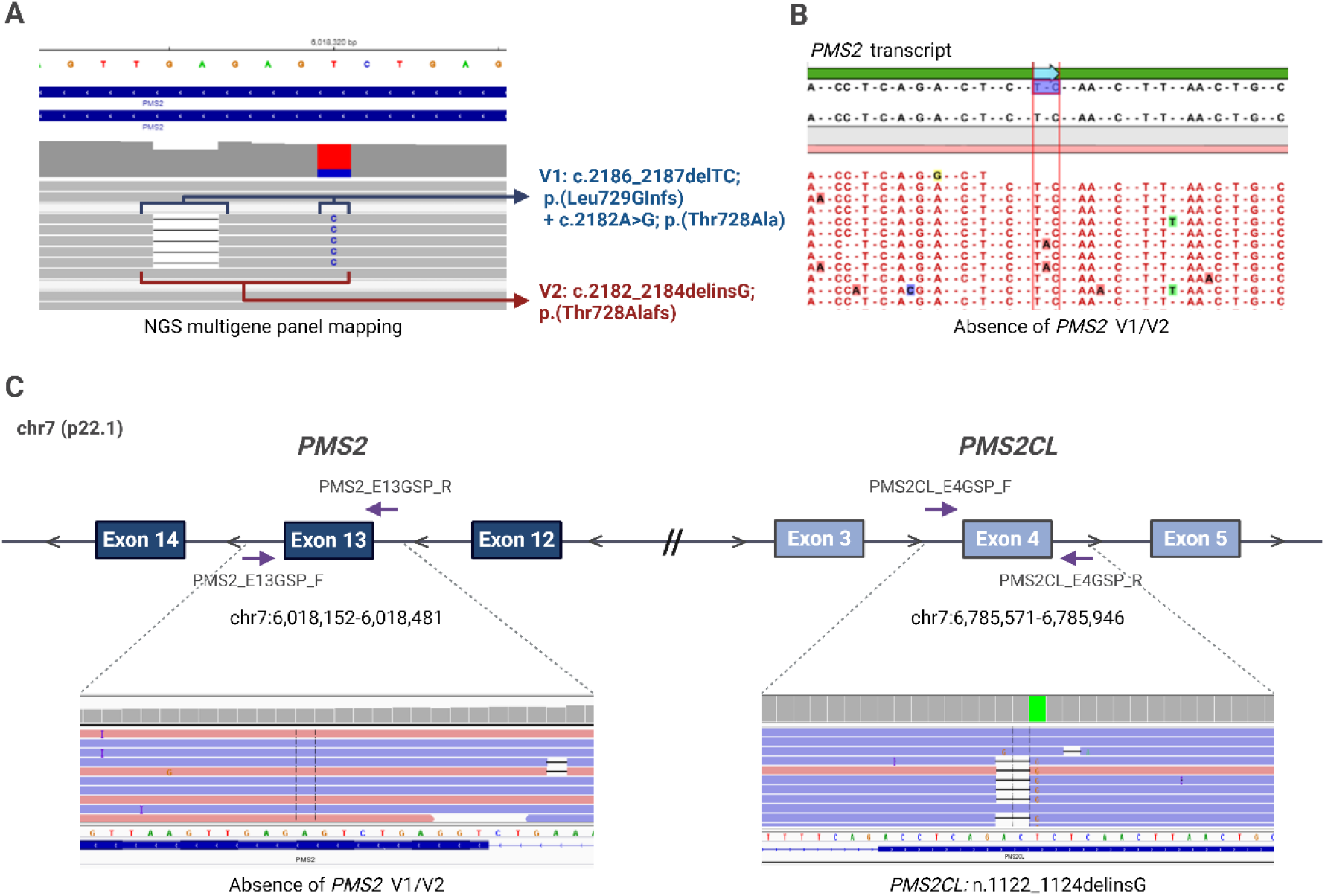
Analysis of *PMS2* V1/V2 variants and reclassification as a *PSM2CL* variant. **A**. NGS reads alignment of multigene DNA panel sequencing, showing the *PMS2* exon 13 region in IGV. The image depicts the bases involved in V1 and V2 nomenclatures. **B**. RNA sequencing for the *PMS2* transcript and analysis by CLC Genomics software, showing absence of V1/V2. **C**. Depiction of gene-specific PCR (GSP) performed for *PM*S2 exon 13 and *PMS2CL* exon 4. Genomic coordinates (Hg19) correspond to amplicons location. Left alignment shows mapping of *PMS2* GSP, demonstrating the absence of V1/V2, and right alignment shows mapping of *PMS2CL* GSP, demonstrating the presence of *PMS2CL*: n.1122_1124delinsG.

Among the 647 individuals evaluated, 12 (1.8%) patients carried the V1/V2 variant, exhibiting variant allele frequencies ranging from 15% to 34% (Table 1). These patients presented distinct cancer types, such as breast (8), colorectal (2) and gastric (1). Four patients had germline pathogenic variants detected in other cancer predisposing genes. The racial distribution of these patients was diverse, with three patients self-identified as Black (one having molecularly confirmed African ancestry), one as Brown and three as White.

**Table 1.** Clinical and genetic features of patients with V1/V2 variant.

| ID | Tumor (age range) | Self-declared race | Genetic testing date | Prior nomenclature | Revised nomenclature | VAF | Additional tests/results |
| --- | --- | --- | --- | --- | --- | --- | --- |
| P_01 | Breast (25-30) | Black | 2018 | PMS2:<br>c.2186_2187delTC | PMS2CL:<br>n.1122_1124delinsG | 25.4% | Ancestry: AFR 66%<sup>a</sup> |
| P_02 | Breast (40-45) | White | 2018 | PMS2:<br>c.2186_2187delTC | PMS2CL:<br>n.1122_1124delinsG | 15.4% | *BRCA1* GPV |
| P_03 | Breast (40-45) | White | 2018 | PMS2:<br>c.2186_2187delTC | PMS2CL:<br>n.1122_1124delinsG | 17.2% | *BRCA1* GPV |
| P_04 | Breast (35-40) | Black | 2018 | PMS2:<br>c.2186_2187delTC | PMS2CL:<br>n.1122_1124delinsG | 34.2% | - |
| P_05 | Gastric (45-0) | White | 2019 | PMS2:<br>c.2186_2187delTC | PMS2CL:<br>n.1122_1124delinsG | 19.4% | - |
| P_06 | CRC (25-30) | NA | 2020 | PMS2:<br>c.2182_2184delinsG | PMS2CL:<br>n.1122_1124delinsG | 20.0% | dMMR (MLH1/PMS2);<br>somatic *MLH1* LoF variant |
| P_07 | Breast (50-55) | NA | 2021 | PMS2:<br>c.2182_2184delinsG | PMS2CL:<br>n.1122_1124delinsG | 14.8% | - |
| P_08 | Rectum (45-50) | Brown | 2022 | PMS2:<br>c.2182_2184delinsG | PMS2CL:<br>n.1122_1124delinsG | 18.4% | *MSH2* GPV |
| P_09 | Breast (50-55) | NA | 2022 | PMS2:<br>c.2182_2184delinsG | PMS2CL:<br>n.1122_1124delinsG | 18.8% | - |
| P_10 | Asymptomatic (40-45) | NA | 2022 | PMS2:<br>c.2182_2184delinsG | PMS2CL:<br>n.1122_1124delinsG | 18.8% | - |
| P_11 | Breast (60-65) | NA | 2023 | PMS2:<br>c.2182_2184delinsG | PMS2CL:<br>n.1122_1124delinsG | 19.0% | *MSH2* GPV |
| P_12 | Breast (60-65) | Black | 2023 | PMS2:<br>c.2182_2184delinsG | PMS2CL:<br>n.1122_1124delinsG | 21.9% | - |

Abbreviations: CRC, colorectal cancer; NA, not available; AFR, African; GPV, germline pathogenic variant; dMMR, deficient mismatch repair; LoF, loss of function. <sup>a</sup>Patient with confirmed African ancestry by Axiom Precision Medicine Diversity Array (PMDA).

We performed gene-specific PCR of all 12 cases and determined that V1/V2 is not present in the *PMS2* gene. Moreover, sequencing analysis of exon 4 of the *PMS2CL* pseudogene unequivocally demonstrated the presence of this variant (Figure 1B). The corresponding correct nomenclature for this variant in *PMS2CL* is n.1122_1124delinsG. To further confirm that V1/V2 is not present in the *PMS2* gene, we conducted an analysis of *PMS2* transcripts by nested PCR in two patients, and in both cases no read containing V1/V2 variant was detected (Figure 1C).

## Discussion

Here, we present data on a frequent *PMS2CL* African variant found in the Brazilian population, which is incorrectly classified as a *PMS2* variant. We detected this variant with two distinct nomenclatures (*PMS2*:c.2182_2184delACTinsG and c.2186_2187delTC) in 1.8% of genetic tests performed in our center. Through gene-specific PCR and *PMS2* transcript analysis, we established that both variants represent the same *PMS2CL* variant (n.1122_1124delinsG). Moreover, in our center 4 patients have received a genetic test report from external laboratories containing this variant mistakenly described as a *PMS2* LoF variant, 3 of them describing the variant as VUS and one as pathogenic, highlighting the relevance of our investigation.

Currently, for the *PMS2* gene, V1: c.2186_2187delTC is classified as “Uncertain significance reviewed by expert panel” and V2 as “Conflicting” in ClinVar, with several laboratories classifying both as “pathogenic.” The conflicting data for classifying both variants is related to the need of confirming the variant as a *PMS2* variant. The annotation by the International Society for Gastrointestinal Hereditary Tumours (InSiGHT) expert group in Clinvar regarding this variant is “This variant is likely to come from pseudogene”. When no confirmation is performed, most likely this variant should be assigned to the *PMS2CL* pseudogene, making it not disease-causing [7].

The V1:c.2186_2187delTC has been reported in the literature in more than 12 articles, published between 1995 and 2019. Few articles described the variant in compound heterozygosity with another *PMS2* variant in patients affected with Turcot Syndrome [8] or Constitutional Mismatch Repair Deficiencies (CMMRD) [9], indicating a true occurrence of the variant in *PMS2*. However, V1 has also been reported in individuals with other cancer types, including colorectal [1, 10], breast [11, 12] and prostate [13], and pseudogene interference was not ruled out in most of these studies. A recent pediatric study reported the finding of V1 in 2 patients with pilocytic astrocytoma, but after applying long-range PCR, they determined that the variant belonged to the pseudogene [14].

The V2: c.2182_2184delinsG is much less cited in the literature, with only 2 articles referring to the variant [1, 5]. The more recent report of this variant most likely reflects updates in variant calling algorithms that incorporated the *PMS2*:c.2182A>G SNV as part of the indel variant, as recommended by HGVS rules. While Guindalini [1] did not describe any sequential technique confirming the variant to *PMS2*, Chong et al showed that indeed the variant was located at *PMS2CL* in all 5 tested patients. In Clinvar, the variant is described as “Conflicting”, with 4 clinical laboratories classifying the variant as Pathogenic/Likely pathogenic and one as VUS. Based on the evidence outlined above, we believe that both V1 and V2 have been incorrectly assigned to *PMS2* and mistakenly classified as pathogenic in several articles and that these variants should be classified as pathogenic only when unequivocal confirmed to be within the *PMS2* gene.

Global genomic databases (ExAC and gnomAD) report an incidence of these variants ranging from 2.5% to 5.3% in the African population, while in ABraOM (a Brazilian genomic database) [15] V1 appears at a frequency of 1.3%. Brazil has a significant percentage of Afro-descendant population with an average African genetic ancestry of 12.7% [5, 16]. In our study, 57% (4/7) of the patients tested with the presence of the *PMS2CL* variant declared themselves as Black or Brown, and one had a 66% African genetic ancestry molecularly confirmed.

Genomics disparities represent a current challenge in clinical genetics [17]. It is well-documented that minorities and underrepresented populations often have more VUS detected in clinical genetic testing. Our report emphasizes this issue by highlighting a common African variant frequently misclassified as a VUS or as pathogenic. Our results also underscore the importance of addressing these misclassification errors caused by NGS’s inability to differentiate short regions that are very similar between *PMS2* and its pseudogenes. The misclassification of variants in the *PMS2* gene can lead to significant consequences in clinical genetics, with clinicians misinterpreting the genetic risk and potentially providing inappropriate management or surveillance recommendations. Moreover, the racial disparity in genomic analysis further exacerbates the challenges faced by minority populations in receiving accurate and timely genetic testing results, stressing the need for increasing the available genomic data from diverse populations.

## Data Availability

All data produced in the present study are available upon reasonable request to the authors.

